# Direct and Indirect Genetic Effects of Birthweight Predisposition on Child DNA Methylation at Birth

**DOI:** 10.1101/2025.06.24.25330182

**Authors:** Nicole Creasey, Leonard Frach, Elena Isaevska, Janine F Felix, Charlotte Cecil, Jean-Baptiste Pingault, Alexander Neumann

## Abstract

**Background:** Birthweight is heritable and strongly associated with epigenetic differences at birth. It is unclear whether a genetic birthweight score is associated with DNA methylation (DNAm) and, if so, whether through direct genetic effects (i.e., child genotype) or indirect genetic effects (i.e., parental genotype, independent of child genotype), which are suspected to be mediated by the prenatal environment (e.g. metabolic factors).

**Methods:** We constructed polygenic scores (PGS) for birthweight predisposition in mothers and children using an existing ‘pre-adjusted’ GWAS assessing maternal indirect effects (maternal genotype on offspring birthweight, correcting for offspring genotype) or fetal effects (offspring genotype on offspring birthweight, correcting for maternal genotype) in 2116 families from the Generation R Study. Offspring DNAm levels at 829 birthweight-related sites were then regressed on both maternal and fetal effect PGS. Additionally, we aggregated 829 DNAm sites into a methylation profile score for birthweight (MPS-BW) and tested which pregnancy health factors (n=13) might mediate genetic effects.

**Findings:** We identified six DNAm sites associated with maternal indirect effects and the birthweight MPS revealed indirect genetic associations, as well. Gestational age partly mediated maternal indirect effects on DNAm, but no other mediators were identified. Results did not depend on offspring sex.

**Interpretation:** This study presents first evidence of maternal genetic effects on offspring birthweight being associated with offspring epigenetic patterns at birth. Paternal genotype was not associated with offspring methylation in this study but was limited by lower sample size and lack of existing GWAS on paternal indirect effects.

**Funding:** European Union

**Research in context:** *Evidence before this study:* Birthweight is a moderately heritable trait and prior genome-wide association studies have identified multiple genetic variants associated with fetal growth. Genetic variants may impact birthweight via direct and indirect genetic routes of transmission. Indirect genetic effects occur when parental genetics still associate with child outcomes independent of genetic transmission to offspring, suggesting involvement of environmental mechanisms. For example, maternal genetic predispositions may shape the intrauterine environment in ways that accelerate or slow fetal growth. Indirect genetic effect may explain up to 22% of birthweight variance. Additionally, epigenetic modifications, particularly differences in DNA methylation (DNAm) at birth, have been linked to birthweight. Recent studies have identified over 900 CpG sites associated with birthweight. However, it is unknown whether the direct and indirect genetic effects on birthweight are reflected in DNAm variation, as well.

*Added value of this study:* We found that especially indirect maternal genetic effects on offspring birthweight are associated with offspring DNAm patterns at birth. We have linked six DNAm sites with indirect maternal effects in contrast to only one site with direct genetic transmission. A methylation profile score for birthweight associated similarly with direct and indirect effects, with a trend for stronger association with indirect effects.

*Implications of all the available evidence:* The results shed new light on how the interplay between genetics and environment on epigenetics may impact birthweight. This study confirms that maternal genetics partly affect offspring birthweight indirectly, independent of direct transmission. Importantly, our results imply that DNAm levels in cord blood partly reflect these indirect maternal genetic effects. This in turn raises the possibility that DNAm is an important mechanism mediating indirect maternal genetic effects on birthweight, but more research is needed to investigate causality.

## Background

Birthweight is a key predictor of infant survival as well as of physical and mental health outcomes across the lifespan^1–4^. Birthweight represents the end result of growth in utero and, therefore, is a valuable marker for prenatal conditions and later health^5^. Intrauterine growth is influenced by genetic factors^6^ as well as in-utero environmental exposures (e.g., metabolic factors or maternal smoking)^7–9^. However, separating the contributions of genetics and environment to birthweight is not straightforward, due to the presence of gene-environment correlations. For example, smoking behavior is partially under genetic influence; thus child genetic effects on birthweight could partly represent a mother’s predisposition towards smoking during pregnancy, which in turn could environmentally influence child development. This represents an indirect genetic effect, that is, a genetic link which is explained by associations between parental genotypes and child outcomes, independent of genetic transmission to offspring. This is in contrast to direct genetic effects, which represent associations between genotypes and child outcomes as a result of genetic transmission.

It has been estimated that 3% to 22% of phenotypic variance in birthweight is explained by indirect genetic effects compared to direct effects from child genetics^6,10–14^. Although causal evidence is currently limited, one of several suggested mechanisms by which genetic and environmental factors may influence child birthweight is via effects on DNA methylation (DNAm). A meta-analytic epigenome-wide association study (*N* = 8825, 24 cohorts) reported associations between birthweight and DNAm at birth at 914 CpG loci.^15^ Since DNAm patterns can be influenced by both the genetics and environment,^16–18^ the same genetic factors involved in direct and indirect genetic effects on birthweight may also affect DNAm profiles. If that is the case, we would expect associations of children’s DNAm at birth with 1) the child’s own genetic birthweight predisposition independent of their parent’s predisposition (i.e., a direct genetic effect), and 2) the parent’s genetic birthweight predisposition independent of the child’s predisposition (i.e., an indirect genetic effect).

Disentangling the effects of genetic birthweight predisposition on DNA methylation could improve our understanding of how genetic predisposition to important health outcomes affects DNAm profiles at birth, which has not been studied before. To our knowledge, only one study^19^ so far applied a trio design to investigate direct and indirect effects of genetic predisposition to a related phenotype – BMI – alongside schizophrenia, educational attainment, and height on offspring methylation in childhood. The authors reported the presence of indirect, mostly maternal, effects on several DNAm sites. However, given the age examined (mean age of 10 years), it remains unclear whether these indirect genetic effects reflect processes occurring in utero (e.g. prenatal maternal smoking), postnatally (e.g. parenting-related behavior) or other forms of gene-environment correlations, such as social stratification^20^. Leveraging DNAm in cord blood offers the advantage of excluding the postnatal environment.

Beyond questions of timing, it is currently unclear whether indirect genetic effects on birthweight depend on offspring sex. As males are, on average, born heavier than females,^21^ and DNAm patterns show widespread sex differences,^22^ it is plausible that direct and indirect genetic effects on birthweight and associated DNAm profiles may differ depending on offspring sex. It is also currently unclear which environmental factors could mediate indirect genetic effects on birthweight, though pregnancy health factors like maternal metabolic factors and blood pressure are plausible candidates.^23^ Finally, an open question is whether different modeling strategies could impact detection of indirect effects. Typically, direct and indirect effects are estimated by computing polygenic scores for each family member using ‘traditional’ GWAS weights, i.e., reflecting effects between one’s own genotype on one’s own birthweight. By mutually adjusting the resulting PGS in subsequent regression models direct and indirect effects can be a posteriori inferred. An alternative approach to the PGS calculation is to test whether direct and indirect effects on DNAm could be better captured by disentangling these sources of variation a priori, before computing PGS, i.e., on the GWAS level. In other words, PGS weights can alternatively be derived from a GWAS reflecting the effects of maternal genotype on child birthweight, independent of child genotype. While such GWAS are not available for most phenotypes, these independent associations of child and maternal genotype have been estimated for birthweight.^6^

To fill these research gaps, in the current paper we estimated direct and indirect genetic effects of birthweight predisposition on 1) DNAm at birth at individual CpG loci previously shown to associate with birthweight and 2) a DNAm-based profile score for birthweight (MPS-BW), based on a sample of up to 2116 families from the Generation R Study (GenR). Furthermore, we tested interactions with sex, tested mediation of 13 pregnancy health factors, and compared different statistical approaches to derive direct and indirect effects.

## Methods

### Participants

The study sample included participants from GenR, an ongoing population-based prospective cohort study of parents and children from fetal life onwards^24^. A total of 9,778 pregnant mothers were enrolled in the study based on the eligibility criteria that they were residents of Rotterdam, the Netherlands, with a delivery date between April 2002 and January 2006. The study was approved by the Medical Ethical Committee of Erasmus MC, University Medical Center Rotterdam, and consent was provided by all families. For the current study, we included 2116 families of European genetic ancestry with complete information on child DNAm measured in cord blood at birth, child and maternal genotype data, and birthweight. Primary analyses were based on these 2116 mother-child duos. However, genotype data of the biological father were also available for 1504 of these families, which were included in supplementary trio analysis. We excluded one sibling per sibling pair based on completeness of data (or at random if equal) and all twin pairs.

### Measures Polygenic scores Genotyping

Genotyping methods have previously been described in detail.^25,26^ Briefly, children were genotyped using Illumina HumanHap 610 or 660 Quad chips, as well as GSA-MD v2.0. Parents were either genotyped with GSA-MD v2.0 or GSA-MD v3.0. Quality checks included Hardy-Weinberg equilibrium, call rate, excess heterozygosity, sex mismatches or relatedness. Imputation was performed using the 1000 Genomes Project (Phase 3, Version 5).^27^ We only retained autosomal SNPs with excellent imputation quality (R² ≥ 0.80) and a minor allele frequency of at least 1% for PGS calculations.

#### GWAS summary statistics and PGS computation

For the main analysis (i.e., mother—child duo analysis) and first supplementary analysis (i.e., trio analysis), PGSs were calculated based on summary statistics from a GWAS of one’s own genotype on own birthweight in a European sample (PGS_child_, PGS_mother_ and PGS_father_)^28^. For the second supplementary analyses (i.e., a priori GWAS adjusted analysis), we used summary statistics from a weighted linear model GWAS in a European sample of 1) maternal genotype on child birthweight adjusted for child genotype to compute a maternal effect PGS (PGS_maternal-effect_) and 2) child genotype on child birthweight adjusted for maternal genotype for a fetal effect PGS (PGS_fetal-effect_)^28^.

Summary statistics to compute the PGS were retrieved from the Early Growth Genetics Consortium website: http://egg-consortium.org/birth-weight-2019.html.^6^ The traditional PGS were based on summary set 1: European-only meta-analysis of own birthweight (298,148 individuals). PGS_fetal-effect_ was based on summary set 5: European-only meta-analysis of the fetal effect; and PGS_maternal-effect_ was based on summary set 6: European-only meta-analysis of the maternal effect in 297,356 individuals with their own birthweight and 210,248 individuals with offspring birthweight (101,541 individuals had both their own and their offspring’s birthweight).

All PGSs were computed with the LDPred2 R package^29^ from child and parent genotype data. We applied automatic tuning of the hyperparameters using ldpred2-auto. To standardize the PGS units and limit batch effects and population stratification, the first ten genetic principal components were regressed on each corresponding z-scored PGS for each family member, and the residuals were used in further analyses.

### DNA Methylation

DNA methylation was quantified with either the Illumina 450k array (GenR_450K_, n=1198) or Illumina EPIC array (GenR_EPIC_, n=918) from genomic DNA extracted from cord blood. To reduce the multiple testing burden and limit both type I and II errors, we extracted probes shown to be associated with child birthweight in a previous meta-EWAS with genome-wide significance^15^, of which 829 of the 914 probes were available in both the GenR_450K_ and GenR_EPIC_ subcohorts. DNAm beta values were converted to M-values to reduce non-normality and improve statistical validity in the analyses^42^. However, to aid interpretation of the effect sizes, the coefficients and confidence intervals based on beta-values were used to visualize the results and are also provided in the supplementary tables. To compute the MPS-BW, the product of the methylation beta value in the current dataset and the corresponding meta-EWAS regression weight was calculated for each CpG, then summed and z-scored to standardize across arrays.

Preparation and normalization of the HumanMethylation450 BeadChip array data (GenR_450k_) and MethylationEPIC v1.0 BeadChip (GenR_EPIC_) was performed according to the CPACOR workflow^30^ with R^31^. Arrays with a call rate above 95% for GenR_450k_ and 96% for GenR_EPIC_ array were retained. Samples with technical failures were removed (i.e., failed bisulfite conversion, hybridization, or extension problems, or mismatched sex determined by the X and Y chromosome probe intensities). The GenR_450k_ data had missing DNAm values for some participants, which were imputed with nearest neighbor averaging using the *impute* package in R^32^. Cell type correction was applied using the cord blood-specific Gervin and Salas method^33^. This method estimates the relative proportions of six white blood cell subtypes (CD4+ T-lymphocytes, CD8+ T-lymphocytes, natural killer cells, B-lymphocytes, monocytes, granulocytes) and nucleated red blood cells (nRBCs). All except nRBCs were included in the statistical models to avoid multi-collinearity.

### Birthweight

Birthweight (in grams) of the children was derived from medical records completed by midwives and obstetricians.

### Potential Mediators

We tested 13 different prenatal health and socioeconomic factors as potential mediators of indirect and direct genetic effects. We selected these factors because they are known determinants of birthweight and have been measured prenatally in the Generation R Study.

Gestational age was based on last menstrual period (LMP) or ultrasound, if LMP could not reliably used for gestational age estimation.^34^ Parity was ascertained via questionnaire. BMI during the first trimester was based on measured weight and height. Presence of gestational hypertensive disorders was extracted from hospital charts (gestational hypertension or (pre-)eclampsia).^35^ Non-fasting insulin levels, folate and homocysteine level were measured before 18 months of gestation in blood plasma.^36,37^ A diet score reflecting adherence to recommended nutrient intakes during the same pregnancy period was computed using a food frequency questionnaire.^38,39^ General maternal psychiatric problems were quantified using the global severity index assessed with the brief symptom inventory.^40^ Information on continued smoking during pregnancy, education and income (once in early pregnancy) was obtained via questionnaires.

### Statistical Analyses

We first tested indirect and direct genetic effects on observed child birthweight itself, using multiple linear regressions with a mother—child duo model (Model 1), trio model (Model 2), and a-priori GWAS adjusted model (Model 3). Child sex (genetically defined) was included as a covariate in all models, while genetic components were already adjusted for by residualizing the PGS. These regression models provided estimates of the effect of a particular family member’s birthweight PGS (i.e. predisposition) on child birthweight controlling for the birthweight PGS of the other family members included in the model. Specifically, the coefficient for the child PGS was interpreted as the direct genetic effect, while the coefficients for the mother PGS and father PGS were interpreted as the maternal and paternal indirect genetic effects, respectively.

1. *M_duo_*: Outcome ∼ PGS_child_ + PGS_mother_ + Covariates
2. *M_trio_*: Outcome ∼ PGS_child_ + PGS_mother_ + PGS_father_ + Covariates
3. *M_gwas-adjusted_*: Outcome ∼ PGS_fetal-effect_ + PGS_maternal-effect_ + Covariates Additionally, for comparison, we performed a series of single PGS regression models whereby each of the PGS was modeled separately as a predictor of child birthweight (models 4-8).
4. Outcome ∼ PGS_child_ + Covariates
5. Outcome ∼ PGS_mother_ + Covariates
6. Outcome ∼ PGS_father_ + Covariates
7. Outcome ∼ PGS_fetal-effect_ + Covariates
8. Outcome ∼ PGS_maternal-effect_ + Covariates

Next, we plotted the coefficients for each family member from the adjusted and unadjusted models with forest plots to allow visual comparison of the estimated effects of birthweight predisposition on child birthweight for each family member with and without adjustment for the birthweight predisposition of other family members.

Furthermore, we tested sex interactions by adding a product term between the PGS and female sex.

Before testing direct and indirect genetic effects on DNAm, we first validated whether the 814 extracted DNAm sites and MPS-BW associate with birthweight in our sample, by regressing birthweight on either a single DNAm site or the MPS-BW. Models were adjusted for covariates child sex, sample plate, and cell type composition. After validation, we then repeated models 1-8 with either DNAm or MPS-BW as an outcome and child sex, sample plate, and cell type composition as covariates. These models allowed us to test the primary research question whether birthweight genetic scores are associated with DNAm at birth via direct or indirect routes of transmission.

Analyses were conducted in R version 4.3.1^31^. An alpha level of .05 was used to test statistical significance and a Benjamini-Hochberg FDR correction^41^ was applied to adjust for multiple testing of 829 DNA methylation sites. Models were fitted separately in the GenR_450K_ and GenR_EPIC_ subcohorts and the results were pooled using inverse-variance based fixed-effect meta-analysis using METAL software^45^ (individual CpG analyses) or the Metafor R package^46^ (MPS and sex-interaction analyses).

To test for mediation, we applied a multiple mediation model in which we assumed that the fetal and maternal effects are causing the outcomes via 13 prenatal factors. Outcomes here refer to either birthweight, DNAm sites showing significant direct or indirect genetic associations or the MPS-BW. Mediation was estimated using a structural equation model with the WLSMV estimator to account for the mix of binary and continuous mediators using the R package lavaan 0.6-19.^43^ We adjusted the b and c paths for sex, and DNA methylation was residualized beforehand for batch and cell proportions to avoid convergence problems. Maternal education and income were defined as continuous variable with 6 and 12 levels, respectively. To keep a consistent sample size with the main analyses and reduce selection bias, we imputed the missing data on mediators using a PCA-based approach, specifically FAMD as implemented in missMDA 1.19.^44^ The optimal number of components was estimated using 10-fold cross-validation. We adjusted for multiple testing of all possible mediation pathways (13 mediators of direct and indirect effects for seven DNA methylation sites = 13*2*7 = 182 mediation pathways).

## Results

### Descriptives

Descriptive statistics can be found in Table 1. Pearson and biserial correlations (for dichotomous variables) between all variables are displayed in Figure S1. As expected, PGS_child_ correlated with PGS_mother_ and PGS_father_ at ca. 0.5 (r=0.54 and r=0.46, respectively). PGS_fetal-effect_ and PGS_maternal-effect_ did not correlate (r=0.04), as the child and mother effects were based on already adjusted weights from a GWAS on fetal and maternal effects. To compare a priori and a posteriori adjustment, we correlated PGS_child_ residualized for PGS_mother_ with PGS_fetal-effect_. The residiualization step is analogous to the a posteriori covariate adjustment in the regression model and allows for direct comparison with the a priori adjusted PGS_fetal-effect_. Resdiualized PGS_child_ and PGS_fetal-effect_ correlated substantially (r=0.81). In contrast, PGS_mother_ adjusted for PGS_child_ did not correlate with PGS_maternal-effect_ (r=0.08), suggesting that both approaches capture different indirect effects.

**Table 1.** Descriptive statistics.

| Variable | Unit | n | mean | sd |
| --- | --- | --- | --- | --- |
| *Continuous variables* |  |  |  |  |
| PGS-Child | SD | 2116 | 0.02 | 0.99 |
| PGS-Maternal | SD | 2116 | 0.03 | 0.98 |
| PGS-Paternal | SD | 1504 | 0.00 | 0.97 |
| PGS-Fetal effect | SD | 2116 | 0.01 | 0.98 |
| PGS-Maternal effect | SD | 2116 | 0.01 | 1.01 |
| Birth weight | g | 2116 | 3534.61 | 509.91 |
| Gestational Age | months | 2116 | 40.10 | 1.48 |
| Parity | Count | 2112 | 0.49 | 0.71 |
| Pregnancy BMI | kg/m<sup>2</sup> | 2057 | 24.61 | 3.94 |
| Maternal age | years | 2116 | 31.97 | 4.22 |
| Insulin | pmol/l | 1739 | 146.03 | 134.57 |
| Folate | nmol/l | 1697 | 20.08 | 8.89 |
| Homocysteine | umol/l | 1678 | 7.22 | 1.87 |
| Diet Score | Score | 1892 | 7.97 | 1.51 |
| Mat. Psychopathology | Score | 1882 | 0.20 | 0.26 |
| cg01899318 | Beta | 2116 | 0.77 | 0.04 |
| cg08817867 | Beta | 2116 | 0.44 | 0.07 |
| cg09119776 | Beta | 2116 | 0.45 | 0.06 |
| cg13182599 | Beta | 2116 | 0.34 | 0.07 |
| cg21707172 | Beta | 2116 | 0.48 | 0.05 |
| cg23663547 | Beta | 2116 | 0.37 | 0.05 |
| cg24020157 | Beta | 2116 | 0.50 | 0.08 |
| Bcell | Proportion | 2116 | 0.06 | 0.02 |
| CD4T | Proportion | 2116 | 0.13 | 0.06 |
| CD8T | Proportion | 2116 | 0.04 | 0.03 |
| Gran | Proportion | 2116 | 0.51 | 0.10 |
| Mono | Proportion | 2116 | 0.09 | 0.03 |
| NK | Proportion | 2116 | 0.07 | 0.03 |
| nRBC | Proportion | 2116 | 0.13 | 0.08 |
| *Categorical variables* | Category | n | n<sub>category</sub> | % |
| Child sex | Female | 2116 | 1065 | 50.3 |
| Education | Higher | 2074 | 1305 | 62.9 |
| Household Income | >2000 Euro | 1929 | 1596 | 82.7 |
| Maternal smoking | Continued smoking | 1962 | 503 | 25.6 |
| Blood pressure complications | Diagnosis | 2102 | 160 | 7.6 |

### Phenotypic analyses: Genetic and DNA methylation effects on birthweight

The results of the preliminary analyses are summarized in Figure 1 and the full model results are shown in Table S1. In the mother-child duo model (M_duo_), a 1SD increase in PGS_child_ significantly associated with a 231.3g increase in birthweight (*b* = 231.3, 95% CI [208.4, 254.3], *p* < .001), indicating a direct genetic effect on child birthweight. On the other hand, PGS_mother_ did not significantly independently of PGS_child_ associate with child birthweight (*b* = 18.7, 95% CI [-4.4, 41.8], *p* = .112). Similarly, in the trio model (M_trio_), each unit increase in PGS_child_ significantly associated with a 212.2g increase in child birthweight (*b* = 212.2, 95% CI [178.8, 245.6], *p* < .001; i.e. evidence of direct genetic effect). However, in contrast to the duo model, each unit increase in PGS_mother_ was significantly associated with a 30.5g increase in child birthweight in the trio model (*b* = 30.5, 95% CI [2.0, 59.0], *p* = .036), thus suggesting a small indirect genetic effect of maternal birthweight predisposition on child birthweight. PGS_father_ did not significantly associate with child birthweight (*b* = 23.6, 95% CI [-5.1, 52.3], *p* = .107). In the a-priori GWAS adjusted analyses (M_gwas-adjusted_), each 1SD increase in the PGS_fetal-effect_ was associated with a 192.52g increase in child birthweight (*b* = 192.5, 95% CI [172.2, 212.8], *p* < .001), while each unit increase in the PGS_maternal-effect_ was significantly associated with a 105.3g increase in child birthweight (*b* = 105.8, 95% CI [86.1, 125.6], *p* < .001) – thus indicating both direct and maternal indirect effects of birthweight on child birthweight.

**Figure 1.**
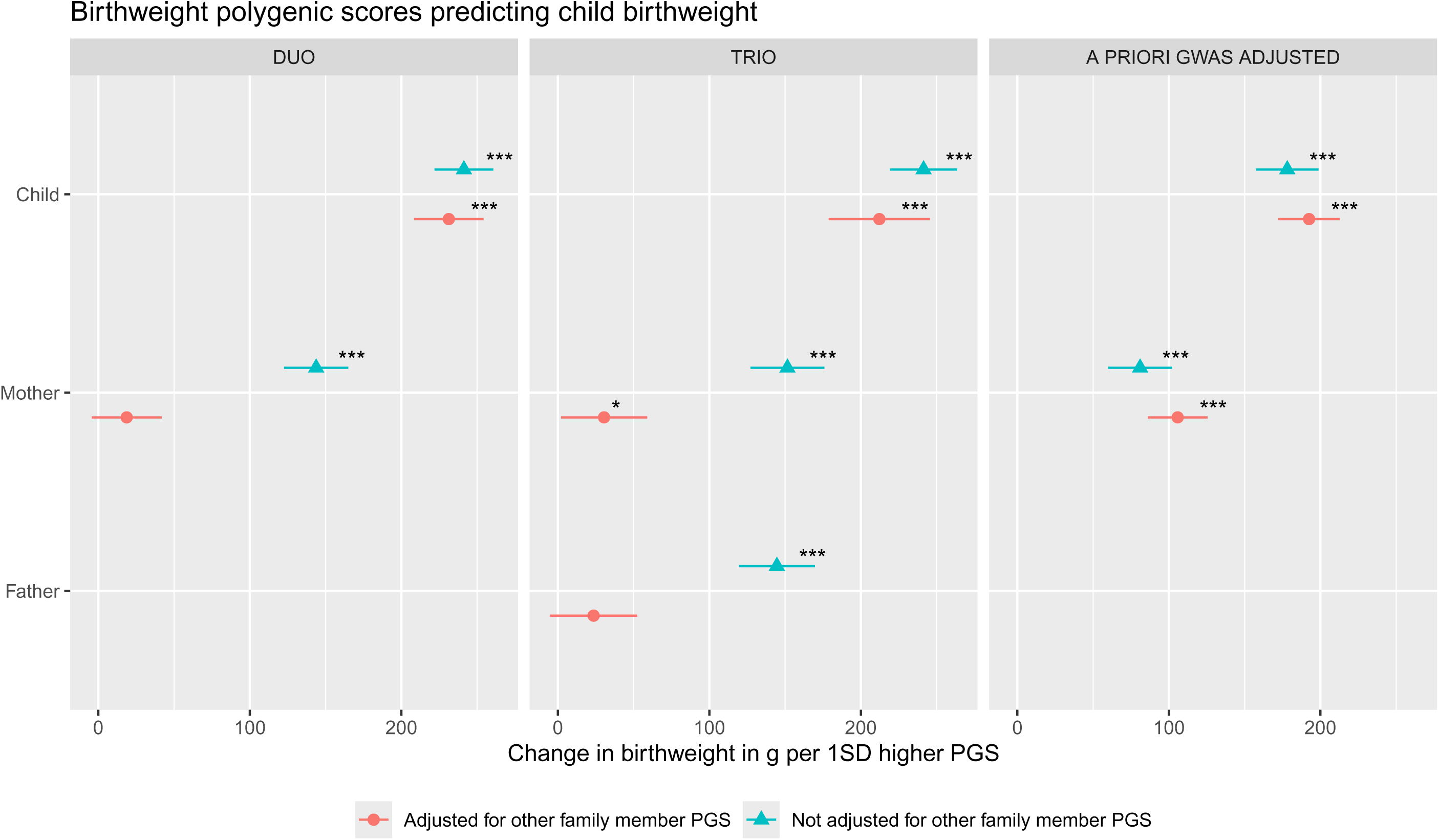
Family polygenic scores predicting child birthweight before and after adjusting for the polygenic scores (PGS) of other family members based on three types of model. In the duo and trio model, PGS were based on GWAS statistics of one’s own genotype on one’s own birthweight in childhood. In the a priori GWAS adjusted model model, PGS were based on a GWAS that already partitions fetal-only genetic effects (PGS_child_) and maternal-only genetic effects (PGS_mother_) on child birthweight. All models were adjusted for child sex and genetic principal components. * p < .05 ** p < .01 *** p < .001

We extended above models by an interaction term between the PGS and child sex, to test whether associations of birthweight genetic predisposition with birthweight and related DNAm patterns differ between males and females. Only the PGS_maternal-effect_ from the a priori-adjusted GWAS (M_gwas-adjusted_) showed suggestive moderation by sex (*b* = 34.71, 95% CI[-4.81, 74.23], *p* = .085, Table S6). The model implies that the maternal genetic indirect effect for lower offspring birthweight is more strongly associated with low birthweight when the child is female (Figure S2).

When testing the associations between the extracted 829 DNAm sites and birthweight, 491 showed FDR-significant associations in the 450K sample (n=1198), 608 in the EPIC sample (n=918) and 639 in the meta-analysis of both (n=2116) (Table S2). A one SD higher MPS-BW was associated with 344.94g (95% CI[317.82, 372.05], *p* < .001) higher birthweight, explaining 16.6% and 23.4% incremental variance beyond covariates alone in the 450K and EPIC subcohorts, respectively (Table S2).

### Main analysis: Genetic effects on birthweight-related DNAm at birth

In the main analysis, we ran mother-child duo regression models (*M_duo_*), this time using DNAm as an outcome instead of birthweight. We examined associations both at the individual CpG level (i.e. with each birthweight-associated CpG tested separately) as well as using the MPS-BW, adjusted for child sex, batch and cell proportions.

In the mother-child duo model (M_duo_), 133 probes showed nominally-significant associations between PGS_child_ and DNAm, independently of PGS_mother_. Of these, one association survived correction for multiple testing (see Table S2 for full results). As shown in Figure 2, PGS_child_ showed a positive association with DNAm at cg23663547 (*b* = 0.02, 95% CI[0.01, 0.03], *p* < .001, *q* = .025), indicating a direct genetic effect of child birthweight predisposition on child DNAm. When aggregating DNAm sites into an MPS-BW, a one SD higher PGS_child_ was associated with 0.08SD (95% CI[0.04, 0.11], *p* < .001) higher MPS-BW (Figure 3).

**Figure 2.**
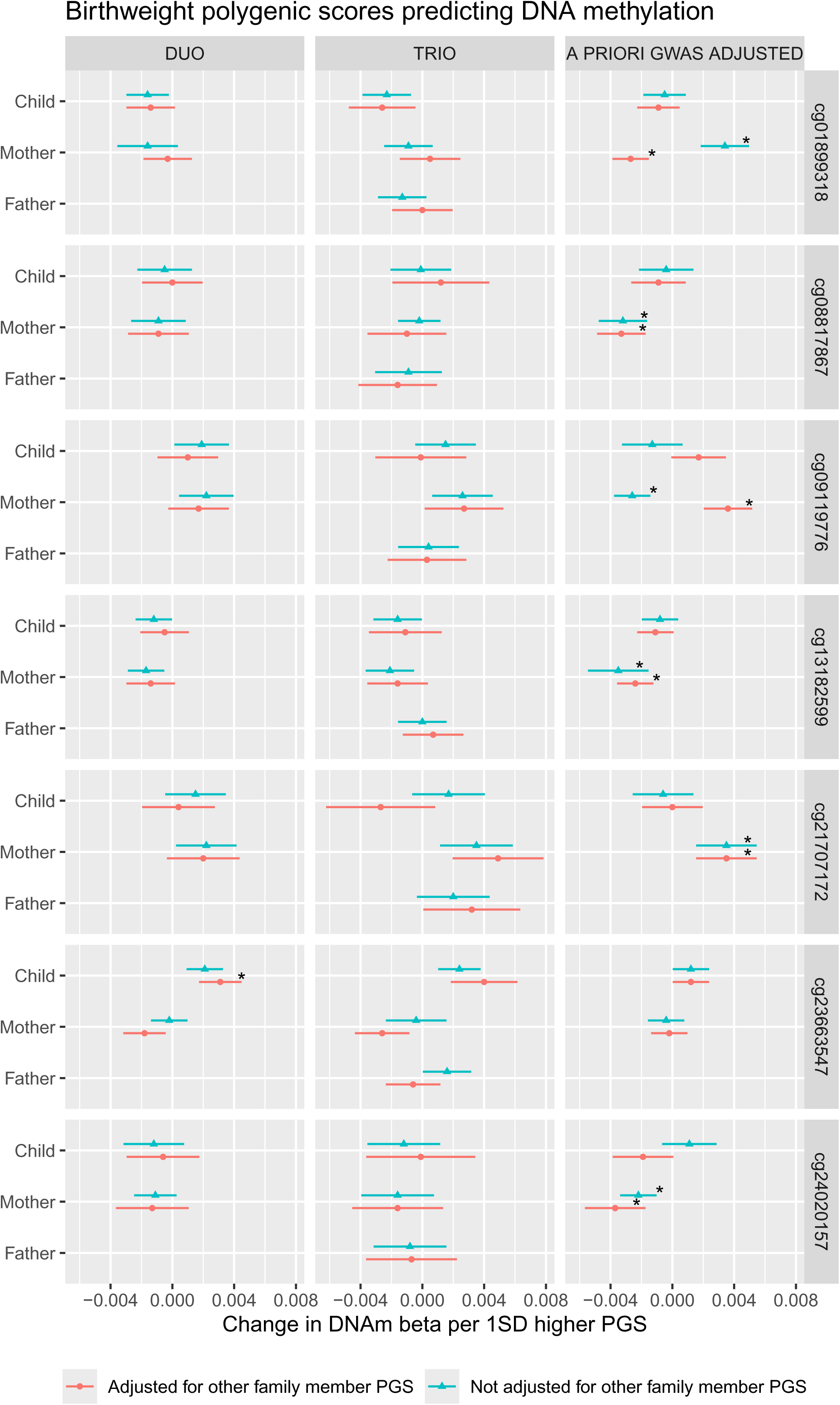
Family polygenic scores predicting DNA methylation at birthweight-associated CpG loci, before and after adjusting for the polygenic scores (PGSs) of other family members based on three types of models. In the duo and trio model, PGSs were based on GWAS statistics of one’s own genotype on one’s own birthweight in childhood. In the a priori GWAS adjusted model, PGS were based on a GWAS that already partitioned fetal-only genetic effects (child PGS) and maternal-only genetic effects (mother PGS) on child birthweight. All models were adjusted for child sex, genetic principal components, DNAm batch and cell proportions. * p < .05 ** p < .01 *** p < .001

**Figure 3.**
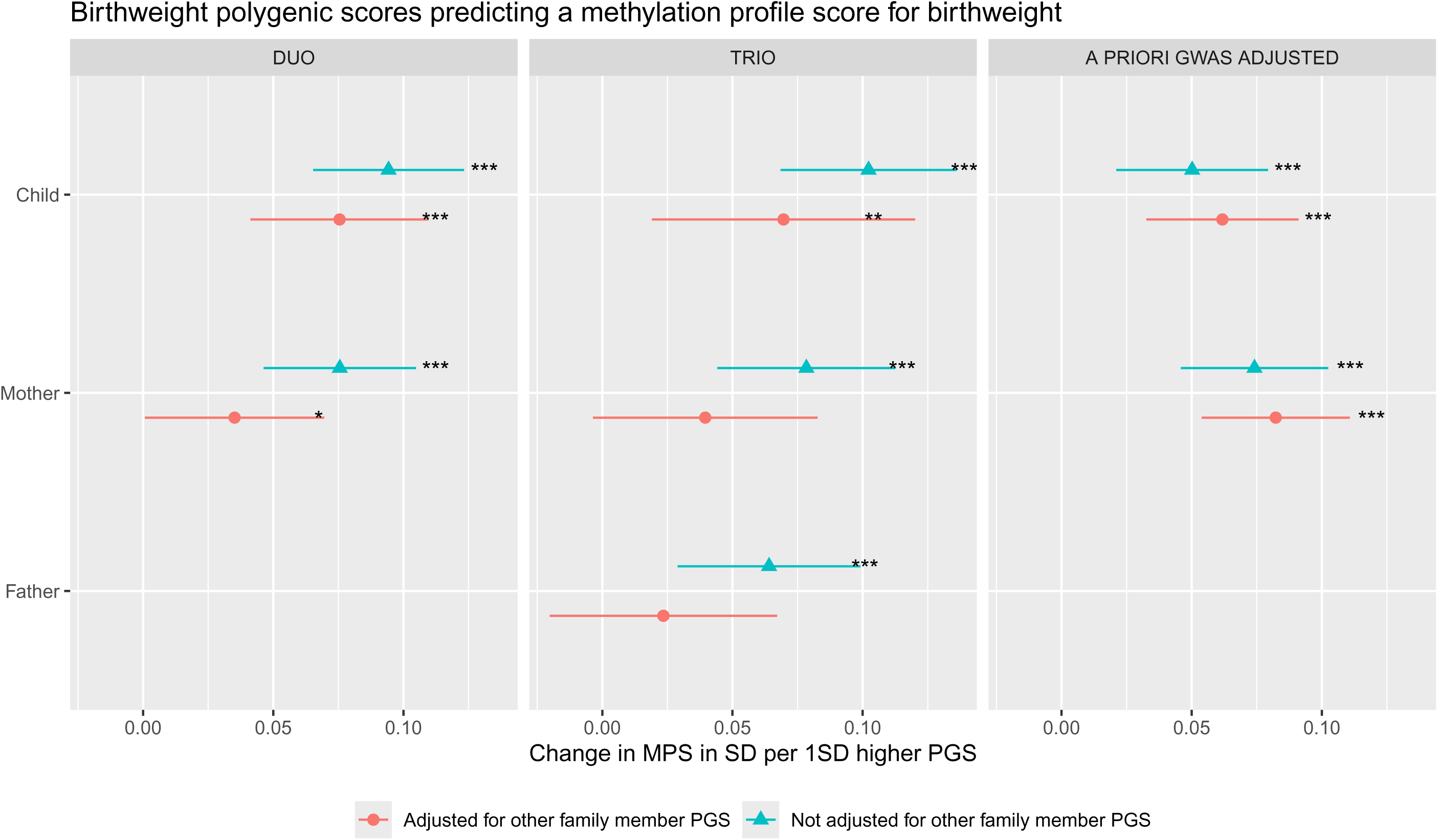
Family polygenic scores predicting a methylation profile score for birthweight, before and after adjusting for the polygenic scores (PGSs) of other family members based on three types of models. In the duo and trio model, PGSs were based on GWAS statistics of one’s own genotype on one’s own birthweight in childhood. In the a priori GWAS adjusted model, PGSs were based on a GWAS that already partitions fetal-only genetic effects (child PGS) and maternal-only genetic effects (mother PGS) on child birthweight. All models were adjusted for child sex, genetic principal components, DNAm batch and cell proportions. * p < .05 ** p < .01 *** p < .001

Maternal PGS showed nominally significant associations with 80 probes, which did not pass correction for multiple testing (Table S3). In contrast, an indirect effect was found for MPS-BW, with a one SD higher PGS_mother_ being associated with 0.04SD (95% CI[0.00, 0.07], *p* < .05) higher MPS-BW.

### Supplementary analyses

#### Trio models

In the first supplementary analysis, we repeated the main analyses also adjusting for father PGS (M_trio_). In the trio models (M_trio_), at nominal significance, 71 probes associated with PGS_child_, 64 probes with PGS_mother_, and 51 probes with PGS_father_. None of the associations survived correction for multiple testing (Table S4). Notably, the FDR-significant effect of PGS_child_ on DNAm at cg23663547 (*CCR7*) identified in the main duo analysis (M_duo_) showed a similar effect size in the trio model and had the smallest p-value (*b* = 0.03, 95% CI[0.01, 0.04], *p* < .001, *q* = .190).

For the MPS-BW, the trio model suggested a similar direct effect as in M_duo_ (*b* = 0.07, 95% CI[0.02, 0.12], *p* = .007). The maternal indirect effect showed also a similar effect size in M_duo_ and M_trio_, but due to higher standard errors in M_trio_ the association was not significant anymore (*b* = 0.04, 95% CI[-0.00, 0.08], *p* = .07). A paternal indirect effect was not significantly associated, either (*b* = 0.02, 95% CI[-0.02, 0.07], *p* = .29) (Table S4, Figure 3).

#### A-priori GWAS adjusted effects models

In the second supplementary analysis, we repeated the main analyses replacing the mother and child PGS with a fetal-effect PGS and maternal-effect PGS (M_gwas-adjusted_). In the a-priori GWAS adjusted models, 108 probes showed nominally significant associations between PGS_fetal-effect_ and DNAm, which did not pass correction for multiple testing (Table S5). In contrast a direct fetal effect was observed for MPS-BW (*b* = 0.06, 95% CI[0.03, 0.09], *p* = <.001).

PGS_maternal-effect_ associated with DNAm at 182 probes, of which 6 associations survived correction for multiple testing as shown in Figure 2, namely: cg01899318 (*ZNF19*), cg08817867, cg09119776 (*SUFU*), cg13182599, cg21707172 (*TIAM2*) and cg24020157 (*RASGEF1A*). DNAm genomic locations can be found in Table S6. PGS_maternal-effect_ also showed a significant association with MPS-BW, notably the effect was comparable to the direct effect, with the point estimate being slightly higher (*b* = 0.08, 95% CI[0.05, 0.11], *p* = <.001).

#### Sex-interaction

Models showing evidence of significant (in/direct) genetic effects on DNAm were repeated to test for sex-interaction by adding a product term between the PGS and female sex. When analyzing birthweight as the outcome, only the PGS_maternal-effect_ showed a trend for sex-interaction. We therefore added to M_gwas-adjusted_ a product term between PGS_maternal-effect_ and sex and tested associations with single DNAm sites, as well as the MPS-BW. No single DNA methylation site nor MPS-BW reached statistical significance for sex interaction after multiple testing correction (p>0.0009, q>0.49, Table S8).

#### Mediation

We further aimed to characterize and understand indirect genetic effects by testing mediation via a number of prenatal health factors and socioeconomic status. Seven DNA methylation sites showed either an indirect (6 sites, M_gwas-adjusted_) or direct (1 site, M_duo_) genetic effect in previous models, with an MPS-BW showing associations with both direct and indirect effects (M_duo,_ M_gwas-adjusted_). As M_gwas-adjusted_ had the most robust findings for indirect effects, we specifically examined whether PGS_maternal-effect_ and PGS_fetal-effect_ associations with birthweight and DNAm were mediated by pregnancy health factors. While each outcome was fitted in separate models, all 13 mediators (gestational age, parity, maternal age, BMI, insulin, blood pressure complications, folate and homocysteine levels, maternal smoking, diet score, maternal psychopathology, maternal education and household income) were included simultaneously and therefore mutually adjusted (see Figure S1 for correlations between PGSs, mediators and outcomes).

The mediation model identified gestational age as a mediator of maternal indirect effects on birthweight and all DNA methylation sites, except for cg01899318. A 1SD higher maternal-effect PGS was associated with 0.12SD higher gestational age, which in turn was associated with 0.46SD higher birthweight. The proportion mediated was 27.6% (95%CI[18.9%; 36.3 %], p<0.001, q<0.001] for birthweight. For the six significant DNA methylation sites the proportion mediated was on average 56% (p<.001, q<.006) and for the MPS the proportion mediated was 26% (95% CI[0.14, 0.37], *p* = <.001), see Table S9. Other pregnancy health factors did not significantly mediate maternal indirect effects and no mediators were identified for fetal effects (p > .15, q=1.00).

## Discussion

This study aimed to better understand how genetic birthweight predispositions influence offspring DNAm at birth by applying a family-based design to separate direct from indirect (parental) routes of transmission. These analyses led to three key findings: First, we identified for the first time indirect genetic effects on DNAm at birth, linking maternal indirect genetic effects for birthweight to methylation levels at six sites. Additionally, one DNAm site was associated with a direct effect. On a broader level, an MPS for birthweight based on 829 DNAm sites was independently associated with both maternal and fetal genetic effects. Second, gestational age partly mediated maternal indirect genetic effects, but not fetal direct effects. Third, computing a maternal PGS using a maternal effect GWAS led to substantially higher estimation of maternal indirect effects than post hoc disentanglement of indirect and direct effects of PGS based on traditional GWAS.Therefore results of the PGS based on a priori adjusted GWAS will be the focus of the discussion.

### Indirect vs direct effects on the methylome

We first tested the genetic effects on birthweight itself. When adjusting child genetic effects for maternal genetics, we observed that most genetic effects on offspring birthweight could be attributed to direct fetal effects, explaining ca. 65% of the total genetic variance, as opposed to 35% attributed to indirect maternal effects. These estimates are somewhat higher than previously reported estimates of 19% based on SNP heritability estimates (M-GCTA) in the MOBA cohort.^6^

While direct effects were more prominent for birthweight, indirect maternal effects were stronger on DNAm levels at birth, based on both single CpG and MPS analyses. For instance, we observed six DNAm sites associated with indirect effects as opposed to only one with direct effects. These sites were located across different chromosomal positions and featured a mix of intergenic and gene-associated CpG regions. All indirect maternal effects were in the expected directions. If a higher genetically predicted birthweight was associated with higher methylation in offspring, then higher methylation was also associated with higher offspring birthweight and vice versa.

This trend of stronger indirect effects appears to generalize to birthweight-associated methylation profiles more broadly, as an MPS for birthweight was also more strongly linked to indirect maternal effects rather than direct fetal effects, though differences were not as pronounced. The results overall suggest that direct genetic effects are more relevant for observed birthweight, but indirect maternal effects have larger or more robust associations for birthweight associated DNAm profiles at birth compared to direct effects. This potentially highlights the sensitivity of DNAm to environmental factors relevant to birthweight. We also tested whether maternal indirect effects depend on the sex of the child, but did not find evidence for such interactions. Finally, we did not see strong evidence for paternal indirect effects, albeit these could not be modeled with an a priori GWAS adjusted PGS.

The identification of indirect maternal effects on DNA methylation raises the question which mechanisms could be at play. A plausible mediator could be the in utero environment: mothers with genetic predisposition to higher offspring birthweight weight may also more likely provide or live in a pregnancy environment conducive to more growth. Our results suggest that birthweight genetic predisposition was not associated with most examined pregnancy factors such as maternal BMI, age, diet, insulin levels, blood pressure complications or smoking behavior. However, gestational age was found to partially mediate the maternal indirect effects on birthweight at most of the tested DNA methylation sites and MPS-BW. Consistent with prior evidence that the maternal indirect effects, but not direct fetal effects, are associated with gestational age,^28^ this suggests that maternal predisposition for higher offspring birthweight may impact the child’s birthweight and DNA methylation by prolonging pregnancy duration. However, because none of the other pregnancy factors mediated the effects, it is difficult to speculate what mechanisms more proximal to the genetic variants may be at play. An alternative interpretation could also be that maternal indirect effects may affect growth, with downstream consequences on length of gestation. The effect direction cannot be disentangled with the current research design and requires future research.

Besides the more proximal gene-environment mechanisms discussed so far, wider gene-environment correlations could also be responsible for the observed associations. For instance, recent evidence suggests that genetic predisposition to educational attainment shows indirect transgenerational effects due to correlations with wider dynastic environments, i.e., the general socioeconomic environment beyond the immediate nuclear family.^20^ Our results suggest that birthweight genetic predisposition was not associated with maternal educational attainment or household income, making a role of socioeconomic effects less likely.

### A priori adjustment of indirect effects

In this study we applied two methods to compute PGS, first using the most common method, i.e. based on a GWAS of one’s own genotype on own phenotype, and second on a GWAS of maternal and fetal effects. When using a traditional PGS, direct and indirect genetic effects are statistically disentangled by calculating the PGS in both parental and offspring genotypes, and modeling these together as predictors in a multiple regression model in order to estimate their independent contribution to offspring birthweight.

An alternative to this post-hoc disentanglement is to calculate PGSs where these effects are already partitioned, i.e. by using weights from a GWAS that already modeled the effect of maternal genotype on offspring birthweight, independent of offspring genotype at a SNP level. This approach has two major advantages compared to the traditional GWAS: 1. Intergenerational effects are explicitly incorporated by estimating the association between parent genotype and child phenotype and 2. Indirect effects are available per SNP. This, in theory, should allow for the computation of a PGS that more comprehensively and more accurately represents indirect effects. These advantages may be especially pronounced for SNPs, which have opposite direct and indirect effects. Indeed, our study results changed substantially when using such an a priori adjusted GWAS to calculate our PGS. While the a posteriori adjusted child PGS correlated strongly with the a priori GWAS-adjusted fetal effect PGS, the two approaches led to very different PGSs for maternal effects, which did not correlate when accounting for child genotype. Furthermore, the maternal-effect PGS based on a priori GWAS adjustment showed 2-3 times higher effect sizes than a posteriori adjustment for the MPS-BW and birthweight. This suggests that post-hoc partitioning of indirect and direct genetic effects at the PGS stage – the most common approach in current research – underestimates the magnitude of indirect genetic effects on birthweight compared to partitioning effects at the GWAS stage, whereas both methods provide similar estimates for direct genetic effects. However, it remains unclear whether such a difference would also hold for other phenotypes since parental effect GWAS are uncommon.

### Strengths and limitations

A major strength of the study was the sophisticated modeling of family genetic effects using modern PGS computation methods and the use of GWAS statistics directly assessing indirect effects a priori. The large sample size and focus on top birthweight associated DNA methylation sites allowed for a statistically powerful analysis with less risk of false positive associations than a hypothesis free approach. At the same time, this focus is also a limitation as it is impossible to draw genome-wide conclusions. For instance, it is unclear whether the pattern of more identified indirect than direct effects would hold when assessing DNA methylation genome-wide beyond the 829 top birthweight associated sites. Furthermore, the power to detect paternal effects was also more limited compared to maternal effects for two reasons: 1. Lower number of paternal genotypes and 2. An a priori adjusted GWAS, which yielded larger indirect effect sizes for mothers, was not available for fathers. Another limitation is that the GenR children participating in this study also contributed to the GWAS of birthweight. However, as they comprised only <0.7% of the total GWAS sample with no overlap in parents, as well as the focus in this study on another phenotype (DNAm) and the partitioning of direct and indirect facts, overfitting is unlikely to explain the results. Finally, while we identified gestational age as a mediator of maternal indirect effects, this is a proximal factor for birthweight. It is unclear what molecular factors more proximal to the maternal genetic variants are mediating the indirect genetic effects.

### Conclusions

In conclusion, this study presents first evidence that maternal indirect genetic effects on offspring birthweight are linked to the offspring methylome. The observed associations between maternal genotype and offspring methylation at six DNA methylation sites could not be explained by direct inheritance but were more compatible with indirect genetic effects. This suggests that birthweight genetic scores are partly a proxy for non-genetic exposures, such as the in utero environment. However, it remains unclear which exact environmental factors are involved, as evidence for mediation via most pregnancy factors or socioeconomic status was weak. The results do, however, suggest that indirect maternal genetic effects may exert effects on DNA methylation through changes in gestational age. Paternal genotype was not associated with offspring methylation in this study but this analysis was limited by a lower sample size and lack of GWAS on paternal indirect effects. Future research should expand analyses to larger genome-wide coverage and include a larger sample of fathers. Finally, more mechanistic research is necessary to understand what pathways are responsible for the observed indirect associations.

## Data Availability

The datasets generated and analyzed during the current study are available upon reasonable request to the head of the Generation R Study.

https://www.generationr.nl

## Acknowledgements

The Generation R Study is conducted by Erasmus MC in close collaboration with the Erasmus University Rotterdam, Faculty of Social Sciences, the Municipal Health Service Rotterdam area, the Rotterdam Homecare Foundation, Rotterdam and the Stichting Trombosedienst & Artsenlaboratorium Rijnmond (STAR-MDC), Rotterdam. The general design of the Generation R Study is made possible by financial support from the Erasmus MC, University Medical Center, Rotterdam, the Netherlands, the Organization for Health Research and Development (ZonMw) and the Ministry of Health, Welfare and Sport. We gratefully acknowledge the contribution of children and their parents, general practitioners, hospitals, midwives and pharmacies in Rotterdam.

This work was supported by the European Union’s HorizonEurope Research and Innovation Programme (FAMILY, grant agreement No 101057529; NC, EI, CAMC, AN; HappyMums, grant agreement No 101057390, CAMC) and the European Research Council (iRISK; grant agreement No 863981; NC, AN, LF, JBP; TEMPO; grant agreement No 101039672; CAMC, AN).

The generation and management of the Illumina 450K and EPIC methylation array data (EWAS data) for the Generation R Study was executed by the Human Genotyping Facility of the Genetic Laboratory of the Department of Internal Medicine, Erasmus MC, the Netherlands. We thank Mr. Michael Verbiest, Ms. Mila Jhamai, Ms. Sarah Higgins, Mr. Marijn Verkerk and Dr. Lisette Stolk for their help in creating the EWAS database. We thank Dr. A.Teumer for his work on the quality control and normalization scripts. The generation and management of GWAS genotype data for the Generation R Study was done at the Human Genomics Facility, HuGe-F, housed within the Laboratory for Population Genomics of the Department of Internal Medicine at Erasmus MC. Genetic Laboratory of the Department of Internal Medicine, Erasmus MC, The Netherlands. We thank Zahra Alawi, Marijn Verkerk, Dr. Katerina Trajanoska, Costanza Vallerga, Samuel Gathan, Dr. Carolina Medina-Gomez, Dr. Linda Broer and Jard de Vries for their help in creating, managing and QC the GWAS database.

## Declerations of interests

The authors declare that they have no competing interests

## Contributors

NC, CAMC, JBP and AN conceptualized the study.

NC and AN performed statistical analysis.

EI contributed to data preparation.

NC, CAMC, JBP and AN wrote the original draft.

JBP and AN supervised the study.

All authors provided feedback on study design and critically revised the manuscript.

## References

1. Knop, M. R. et al. Birth Weight and Risk of Type 2 Diabetes Mellitus, Cardiovascular Disease, and Hypertension in Adults: A Meta-Analysis of 7 646 267 Participants From 135 Studies. JAHA 7, e008870 (2018).

2. McCormack, V. A., Dos Santos Silva, I., Koupil, I., Leon, D. A. & Lithell, H. O. Birth characteristics and adult cancer incidence: Swedish cohort of over 11,000 men and women. Intl Journal of Cancer 115, 611–617 (2005).

3. Lawlor, D. A., Ronalds, G., Clark, H., Davey Smith, G. & Leon, D. A. Birth Weight Is Inversely Associated With Incident Coronary Heart Disease and Stroke Among Individuals Born in the 1950s: Findings From the Aberdeen Children of the 1950s Prospective Cohort Study. Circulation 112, 1414–1418 (2005).

4. Mathewson, K. J. et al. Mental health of extremely low birth weight survivors: A systematic review and meta-analysis. Psychological Bulletin 143, 347–383 (2017).

5. Barker, D. J. P. Fetal origins of coronary heart disease. BMJ 311, 171–174 (1995).

6. Warrington, N. M. et al. Maternal and fetal genetic effects on birth weight and their relevance to cardio-metabolic risk factors. Nat Genet 51, 804–814 (2019).

7. Di, H.-K., et al. Maternal smoking status during pregnancy and low birth weight in offspring: systematic review and meta-analysis of 55 cohort studies published from 1986 to 2020. World J Pediatr 18, 176–185 (2022).

8. Yitshak-Sade, M., et al. Estimating the Combined Effects of Natural and Built Environmental Exposures on Birthweight among Urban Residents in Massachusetts. IJERPH 17, 8805 (2020).

9. Englund-Ögge, L., et al. Associations between maternal dietary patterns and infant birth weight, small and large for gestational age in the Norwegian Mother and Child Cohort Study. Eur J Clin Nutr 73, 1270–1282 (2019).

10. Beaumont, R. N. et al. Genome-wide association study of offspring birth weight in 86 577 women identifies five novel loci and highlights maternal genetic effects that are independent of fetal genetics. Human Molecular Genetics 27, 742–756 (2018).

11. Eaves, L. J., Pourcain, B. St., Smith, G. D., York, T. P. & Evans, D. M. Resolving the Effects of Maternal and Offspring Genotype on Dyadic Outcomes in Genome Wide Complex Trait Analysis (“M-GCTA”). Behav Genet 44, 445–455 (2014).

12. Magnus, P. Causes of variation in birth weight: A study of offspring of twins. Clinical Genetics 25, 15–24 (1984).

13. Magnus, P. Further evidence for a significant effect of fetal genes on variation in birth weight. Clinical Genetics 26, 289–296 (1984).

14. CHARGE Consortium Hematology Working Group, et al. Genome-wide associations for birth weight and correlations with adult disease. Nature 538, 248–252 (2016).

15. Küpers, L. K. et al. Meta-analysis of epigenome-wide association studies in neonates reveals widespread differential DNA methylation associated with birthweight. Nat Commun 10, 1893 (2019).

16. Gaunt, T. R. et al. Systematic identification of genetic influences on methylation across the human life course. Genome Biology 1–14 (2016) doi:10.1186/s13059-016-0926-z.

17. Chatterjee, S., Ouidir, M. & Tekola-Ayele, F. Genetic and in utero environmental contributions to DNA methylation variation in placenta. Human Molecular Genetics 30, 1968–1976 (2021).

18. Joubert, B. R. et al. DNA Methylation in Newborns and Maternal Smoking in Pregnancy: Genome-wide Consortium Meta-analysis. The American Journal of Human Genetics 98, 680–696 (2016).

19. Hagenbeek, F. A. et al. Intergenerational transmission of complex traits and the offspring methylome. 2024.04.15.24305824 Preprint at 10.1101/2024.04.15.24305824 (2024).

20. Nivard, M. G. et al. More than nature and nurture, indirect genetic effects on children’s academic achievement are consequences of dynastic social processes. Nat Hum Behav 8, 771–778 (2024).

21. The WHO Child Growth Standards. https://www.who.int/tools/child-growth-standards.

22. Solomon, O. et al. Meta-analysis of epigenome-wide association studies in newborns and children show widespread sex differences in blood DNA methylation. Mutation Research/Reviews in Mutation Research 789, 108415 (2022).

23. Tyrrell, J. et al. Genetic evidence for causal relationships between maternal obesity-related traits and birth weight. JAMA - Journal of the American Medical Association 315, 1129–1140 (2016).

24. Kooijman, M. N. et al. The Generation R Study: design and cohort update 2017. European Journal of Epidemiology 31, 1243–1264 (2016).

25. Ghatan, S. et al. Genetic Nurture: Estimating the direct genetic effects of pediatric anthropometric traits. 2024.12.10.24318796 Preprint at 10.1101/2024.12.10.24318796 (2024).

26. Medina-Gomez, C., et al. Challenges in conducting genome-wide association studies in highly admixed multi-ethnic populations: the Generation R Study. European journal of epidemiology 30, 317–330 (2015).

27. The 1000 Genomes Project Consortium. A global reference for human genetic variation. Nature 526, 68–74 (2015).

28. EGG Consortium et al. Maternal and fetal genetic effects on birth weight and their relevance to cardio-metabolic risk factors. Nat Genet 51, 804–814 (2019).

29. Privé, F., Arbel, J. & Vilhjálmsson, B. J. LDpred2: better, faster, stronger. Bioinformatics 36, 5424–5431 (2021).

30. Lehne, B. et al. A coherent approach for analysis of the Illumina HumanMethylation450 BeadChip improves data quality and performance in epigenome-wide association studies. Genome Biol 16, 37 (2015).

31. R Core Team. R: A Language and Environment for Statistical Computing. R Foundation for Statistical Computing, Vienna. http://www.R-project.org/. (2013).

32. Hastie, T., Narasimhan, B. & Chu, G. impute: Imputation for microarray data. (2023).

33. Gervin, K. et al. Systematic evaluation and validation of reference and library selection methods for deconvolution of cord blood DNA methylation data. Clin Epigenet 11, 125 (2019).

34. Verburg, B. O. et al. New charts for ultrasound dating of pregnancy and assessment of fetal growth: Longitudinal data from a population-based cohort study. Ultrasound in Obstetrics and Gynecology 31, 388–396 (2008).

35. Benschop, L. et al. Gestational hypertensive disorders and retinal microvasculature: the Generation R Study. BMC Med 15, 153 (2017).

36. Wahab, R. J. et al. Maternal Glucose Concentrations in Early Pregnancy and Cardiometabolic Risk Factors in Childhood. Obesity 28, 985–993 (2020).

37. Monasso, G. S., Küpers, L. K., Jaddoe, V. W. V., Heil, S. G. & Felix, J. F. Associations of circulating folate, vitamin B12 and homocysteine concentrations in early pregnancy and cord blood with epigenetic gestational age: the Generation R Study. Clinical Epigenetics 13, 95 (2021).

38. Klipstein-Grobusch, K., et al. Dietary assessment in the elderly: validation of a semiquantitative food frequency questionnaire. Eur J Clin Nutr 52, 588–596 (1998).

39. Nguyen, A. N. et al. Maternal history of eating disorders: Diet quality during pregnancy and infant feeding. Appetite 109, 108–114 (2017).

40. Derogatis, L. R. & Melisaratos, N. The brief symptom inventory: an introductory report. Psychological medicine 13, 595–605 (1983).

41. Benjamini, Y. & Hochberg, Y. Controlling the false discovery rate: A practical and powerful approach to multiple testing. Journal of the Royal Statistical Society: Series B (Methodological) 57, 289–300 (1995).

42. Du, P. et al. Comparison of Beta-value and M-value methods for quantifying methylation levels by microarray analysis. BMC Bioinformatics 11, 587 (2010).

43. Rosseel, Y. lavaan: An R package for structural equation modeling. Journal of Statistical Software (2012).

44. Josse, J. & Husson, F. missMDA: A package for handling missing values in multivariate data analysis. Journal of Statistical Software 70, (2016).

45. Willer, C. J., Li, Y. & Abecasis, G. R. METAL: fast and efficient meta-analysis of genomewide association scans. Bioinformatics 26, 2190–2191 (2010).

46. Viechtbauer, W. Conducting Meta-Analyses in R with the metafor Package. J. Stat. Soft. 36, (2010).

