## Supplementary figures and images for "Direct and Indirect Genetic Effects of Birthweight Predisposition on Child DNA Methylation at Birth"

### Figure S1

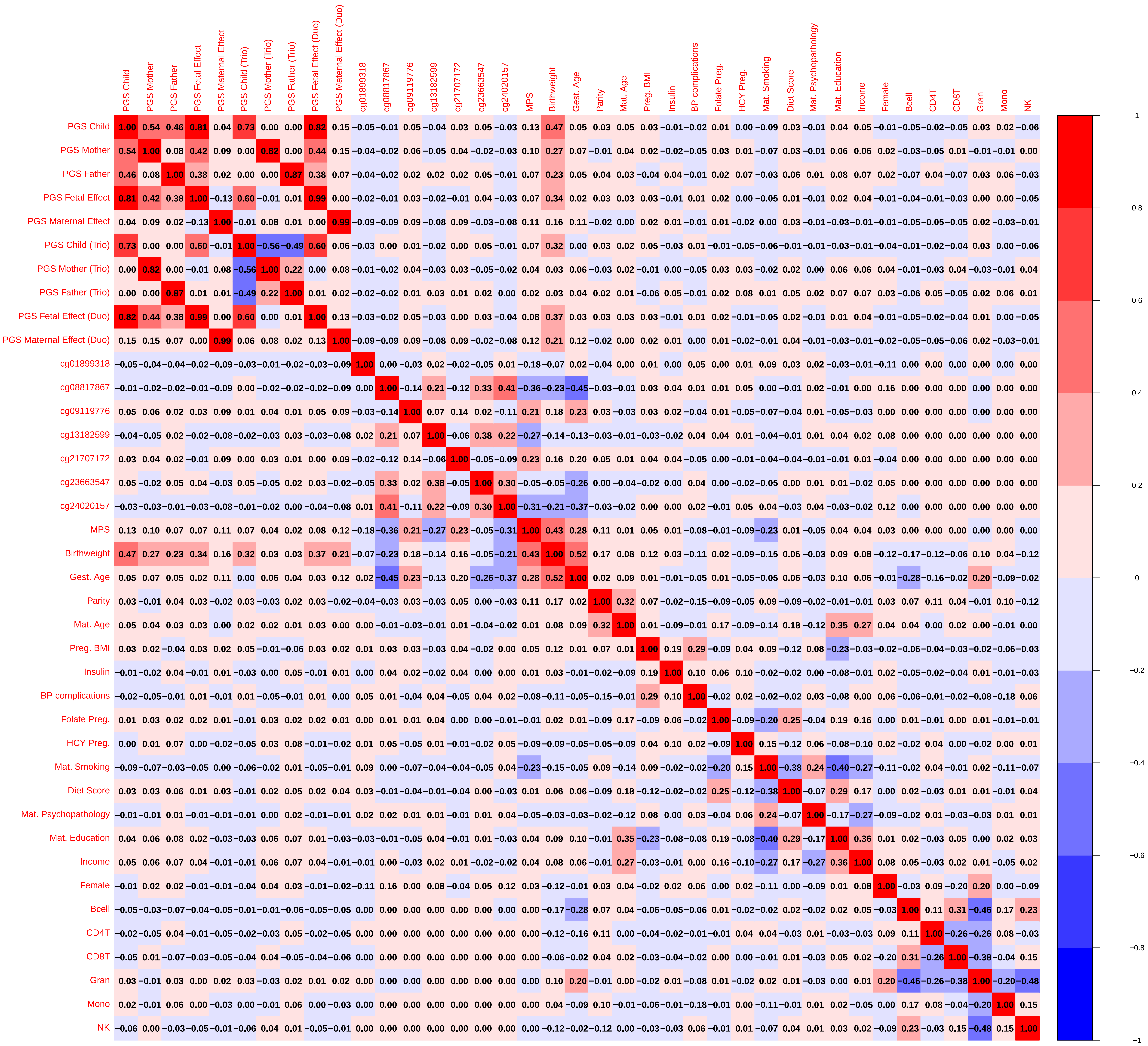

### Figure S2

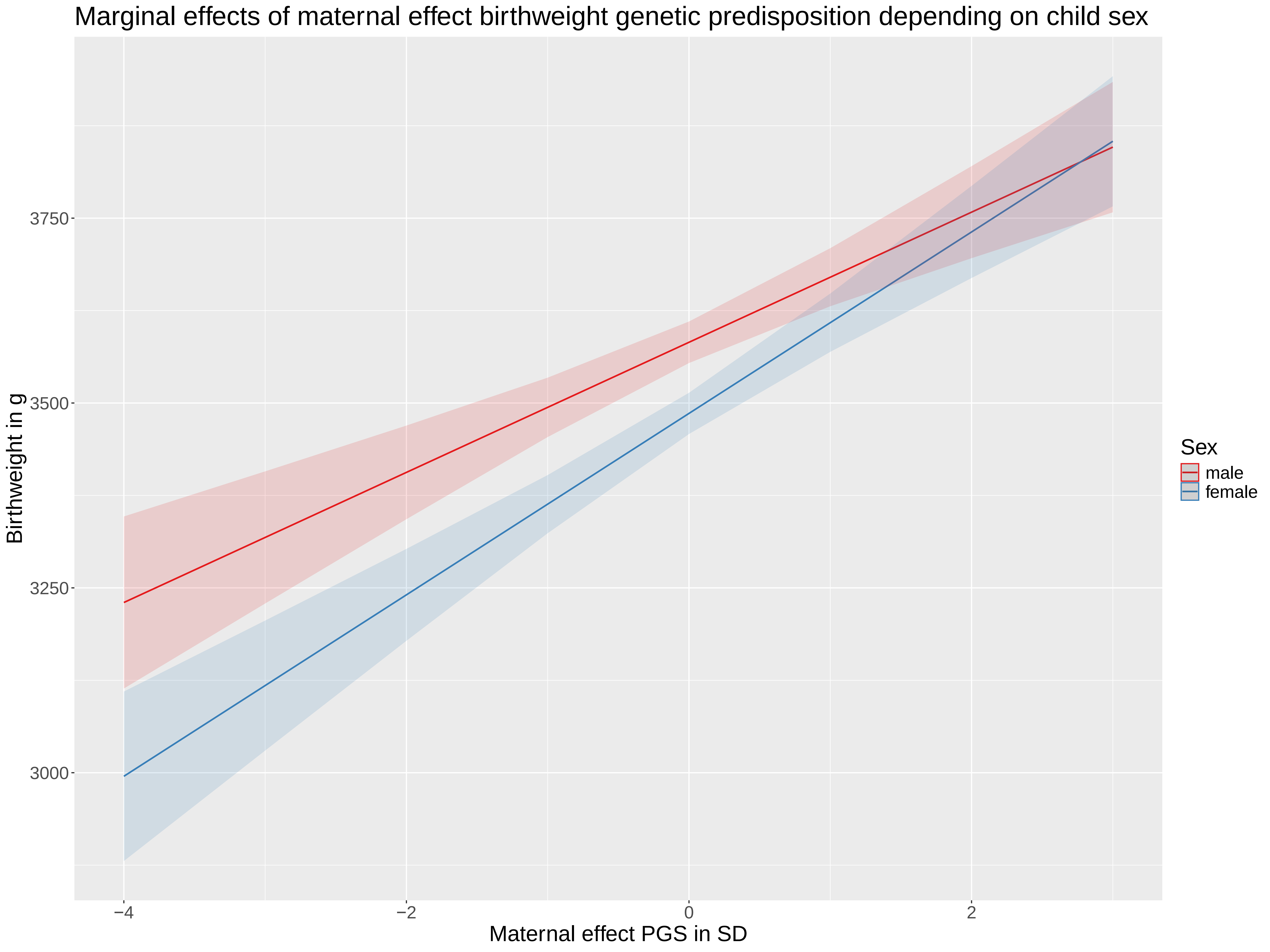
